# The male excess case-fatality rates for COVID-19 – A meta-analytic study of the age-related differences and consistency over six countries

**DOI:** 10.1101/2020.06.11.20128439

**Authors:** Manfred S Green, Naama Schwartz, Dorit Nitzan, Victoria Peer

**Author notes:** **Correspondence:** Prof. Manfred Green MBChB,PhD, School of Public Health, University of Haifa, Abba Koushy 199, Haifa 3498838, Israel. **Availability of data and materials** All data are available from the original sources or from the authors. **Competing interests** The authors declare that they have no competing interests. **Funding** No funding sources were used for the study. **Authors’ contributions** MSG designed and supervised the study and participated in the analysis and interpretation of the data and in writing the manuscript. NS assisted in the data analysis and contributed important input in the review of the manuscript. DN participated in the conception of the study and critically evaluated the manuscript. VP participated in the study design, collected the data, helped in the interpretation of the analyses and writing the manuscript. All authors approved the final version submitted.

## Abstract

**Background:** Early in the COVID-19 pandemic, it was noted that males seemed to be more affected than females. We examined the magnitude and consistency of the sex differences in age-specific case-fatality rates (CFRs) in six countries.

**Methods:** Data on the cases and deaths from COVID-19, by sex and age group, were extracted from the published reports from Denmark, England, Israel, Italy, Spain, and the United States. Age-specific CFRs were computed for males and females separately. The ratio of the male to female CFRs were computed and meta-analytic methods were used to obtained pooled estimates of the male to female ratio of the CFRs over the six countries, for seven age-groups.

**Findings:** The CFRs were consistently higher in males at all ages. The differences were greater in the younger age groups. The pooled M:F CFR ratios were 2.53, 2.92, 2.57, 1.83, 1.57, 1.58 and 1.48 for ages 0-39, 40-49, 50-59, 60-69, 70-79, 80-89 and 90+. There was remarkable consistency between countries in the magnitude of the M:F CFRs, in each age group. In meta-regression, age group explained almost all the heterogeneity in the CFR ratios.

**Conclusions:** The sex differences in the CFRs are intriguing and are compatible with the male dominance in the incidence rates of many infectious diseases. For COVID-19, factors such as sex differences in the prevalence of underlying diseases may play a part in the CFR differences. However, the greater severity of the disease in males, particularly at younger ages, may be part of the disease mechanism and should be explored further.

**Funding:** No funding was provided for this study. The authors declare no conflict of interests

## Background

Early in the course of the COVID-19 pandemic, it was observed that the incidence of the disease appeared to be higher in males (1). However, this observation has been inconsistent and it is possible that some of the differences observed were due to differences in exposure. In some countries, many of the cases were healthcare workers, who may be overrepresented by females (2). In South Korea, a large outbreak of COVID-19 occurred in a religious group whose members were predominantly young women (3). While incidence rates are directly related to exposure, the case-fatality rate (CFR) is a measure of the severity of the disease which is not usually affected by exposure. It has been reported that the CFR is higher in males than females (4, 5).

Sex differences in infectious diseases can provide clues to the mechanisms of infections (6). Males tend to have a higher incidence in many infectious diseases, although not in all age groups (6-8), whereas females dominate in the incidence of diseases such as pertussis (9). The exact mechanism of the sex differences in the incidence and mortality from infectious diseases is not well understood (6). Elucidation of the sex differences in COVID-19 could contribute to a better understanding of the pathogenesis of COVID-19. In this paper we used meta-analytic methods to examine the magnitude and consistency of sex differences in the COVID-19 CFRs by age group, in six countries.

## Methods

### Study design

A meta-analytic study of retrospective national cohorts of COVID-19.

### Source of data

Data on COVID-19 were extracted from published reports from six countries. The number of cases and deaths in each age group by sex were available for Denmark, England, Israel, Italy, Spain and United States. Data on deaths with confirmed COVID-19 infection for Denmark were obtained from published reports of Statens Serum Institut (SSI) (10), for England from Office for National Statistics (ONS) (11). Data on cases with confirmed COVID-19 infection for Denmark and England were extracted from Statista.com (12). Data on cases and deaths for Israel were obtained from the Israeli Ministry of Health database. Data on cases and deaths for Italy were obtained from Istituto Superiore di Sanità (13), for Spain from the Ministerio De Sanidad, Centro de Coordinación de Alertas y Emergencias Sanitarias (14) and for the United States from the published article (15).

### Statistical analyses

COVID-19 CFRs by sex and age group using the number of deaths divided by the number of reported cases. The age groups were divided by intervals: 0-39, 40-49, 50-59, 60-69,70-79, 80-89, 90+. The ratio of the male to female CFRs (CFRR) was calculated by dividing the CFRs for males by the CFRs for females, by age group and country. We used meta-analytic methodology to evaluate the overall magnitude and consistency of the sex differences in the CFR of COVID-19 by age group, across different countries. The outcome variable was the male to female CFRR. The data presented (forest plots) are the CFRRs by age group and country. Heterogeneity was evaluated using Cochran’s Q statistic. Tau^2^ and I^2^ were used to estimate the between-study variance. If the Q test yielded a p < 0.1, and/or I^2^ ≥50%, the random effects model (16) was used to estimate pooled CFRRs and 95% confidence intervals (CI), otherwise the fixed model was used. To evaluate the effect of individual county on the risk of COVID-19, we performed leave-one-out sensitivity analysis and recomputed the pooled CFRRs. We performed the Egger test for asymmetry for testing for a possible imbalance in the studies around the pooled CFRR. In order to explore associations of the CFRs with sex, age-group and country, meta-regression analyses were performed. The meta-analyses and meta-regressions were carried out using STATA software version 12.1 (Stata Corp., College Station, TX).

### Ethics

National, open access aggregative and anonymous data were used and there was no need for ethics committee approval.

## Results

### Descriptive statistics

The summary of male and female CFRs in different countries for each age group is presented in Table 1. The larger number of female cases in many countries may reflect more exposure in the workplace. However, over countries and time periods, the CFRs were consistently higher among males compared with females. Up to the age of 70, the male CFRs are more than double those of the female, and over the age of 70, the male CFRs were 24% to 99% higher.

**Table 1.** Cases, deaths and CFRs by age group and sex

| **Age Group** | **Countries** | **Males cases** | **Males death** | **Male Fatality %** | **Females cases** | **Females death** | **Female Fatality%** | **Male/ Female Case Fatality %** |
| --- | --- | --- | --- | --- | --- | --- | --- | --- |
| 0-39 | England | 9,940 | 125 | 1.26 | 18,124 | 90 | 0.50 | 2.53 |
|  | Israel | 5,275 | 2 | 0.04 | 4,182 | 1 | 0.02 | 1.59 |
|  | Italy | 14,557 | 42 | 0.29 | 17,496 | 24 | 0.14 | 2.10 |
|  | Spain | 13,903 | 58 | 0.42 | 24,644 | 35 | 0.14 | 2.94 |
|  | USA | 186 | 9 | 4.84 | 156 | 3 | 1.92 | 2.52 |
| 40-49 | England | 7,197 | 323 | 4.49 | 9,936 | 185 | 1.86 | 2.41 |
|  | Israel | 1,082 | 2 | 0.18 | 884 | 1 | 0.11 | 1.63 |
|  | Italy | 11,590 | 184 | 1.59 | 15,908 | 62 | 0.39 | 4.07 |
|  | Spain | 14,352 | 131 | 0.91 | 20,474 | 70 | 0.34 | 2.67 |
|  | USA | 233 | 19 | 8.15 | 119 | 3 | 2.52 | 3.23 |
| 50-59 | England | 9,629 | 1,105 | 11.48 | 11,403 | 573 | 5.02 | 2.28 |
|  | Israel | 1,034 | 3 | 0.29 | 870 | 6 | 0.69 | 0.42 |
|  | Italy | 18,047 | 778 | 4.31 | 20,297 | 215 | 1.06 | 4.07 |
|  | Spain | 18,281 | 434 | 2.37 | 24,479 | 177 | 0.72 | 3.28 |
|  | USA | 327 | 40 | 12.23 | 188 | 13 | 6.91 | 1.77 |
| 60-69 | Denmark | 651 | 33 | 5.07 | 638 | 20 | 3.13 | 1.62 |
|  | England | 9,105 | 2,366 | 25.99 | 6,919 | 1,167 | 16.87 | 1.54 |
|  | Israel | 859 | 17 | 1.98 | 595 | 9 | 1.51 | 1.31 |
|  | Italy | 17,678 | 2,299 | 13.00 | 11,529 | 676 | 5.86 | 2.22 |
|  | Spain | 17,828 | 1,198 | 6.72 | 16,506 | 497 | 3.01 | 2.23 |
|  | USA | 300 | 56 | 18.67 | 233 | 28 | 12.02 | 1.55 |
| 70-79 | Denmark | 516 | 110 | 21.32 | 427 | 46 | 10.77 | 1.98 |
|  | England | 11,639 | 5,033 | 43.24 | 8,346 | 2,854 | 34.20 | 1.26 |
|  | Israel | 467 | 43 | 9.21 | 376 | 23 | 6.12 | 1.51 |
|  | Italy | 18,355 | 5,566 | 30.32 | 13,231 | 2,283 | 17.25 | 1.76 |
|  | Spain | 17,889 | 3,165 | 17.69 | 14,533 | 1,468 | 10.10 | 1.75 |
|  | USA | 254 | 91 | 35.83 | 197 | 54 | 27.41 | 1.31 |
| 80-89 | Denmark | 325 | 121 | 37.23 | 431 | 87 | 20.19 | 1.84 |
|  | England | 13,353 | 7,317 | 54.80 | 13,686 | 5,616 | 41.03 | 1.34 |
|  | Israel | 200 | 51 | 25.50 | 236 | 43 | 18.22 | 1.40 |
|  | Italy | 15,784 | 6,593 | 41.77 | 22,206 | 4,801 | 21.62 | 1.93 |
|  | Spain | 15,625 | 4,372 | 27.98 | 21,805 | 3,500 | 16.05 | 1.74 |
|  | USA | 155 | 94 | 60.65 | 158 | 76 | 48.10 | 1.26 |
| 90+ | Denmark | 66 | 37 | 56.06 | 175 | 78 | 44.57 | 1.26 |
|  | England | 4,347 | 2,838 | 65.29 | 7,642 | 3,773 | 49.37 | 1.32 |
|  | Israel | 70 | 29 | 41.43 | 162 | 54 | 33.33 | 1.24 |
|  | Italy | 3,632 | 1,556 | 42.84 | 13,334 | 2,873 | 21.55 | 1.99 |
|  | Spain | 5,105 | 1,589 | 31.13 | 13,195 | 2,493 | 18.89 | 1.65 |
|  | USA | 44 | 28 | 63.64 | 84 | 39 | 46.43 | 1.37 |

### Meta-analyses by age group

The forest plot for the case-fatality rates by age group and country is shown in Figure 1. The highest CFRRs are in the youngest groups and decline with age. The pooled M:F CFR ratios were 2.53, 2.92, 2.57, 1.83, 1.57, 1.58 and 1.48 for ages 0-39, 40-49, 50-59, 60-69, 70-79, 80-89 and 90+. There was remarkable consistency between countries in the magnitude of the M:F CFRs in each age group.

**Figure 1:**
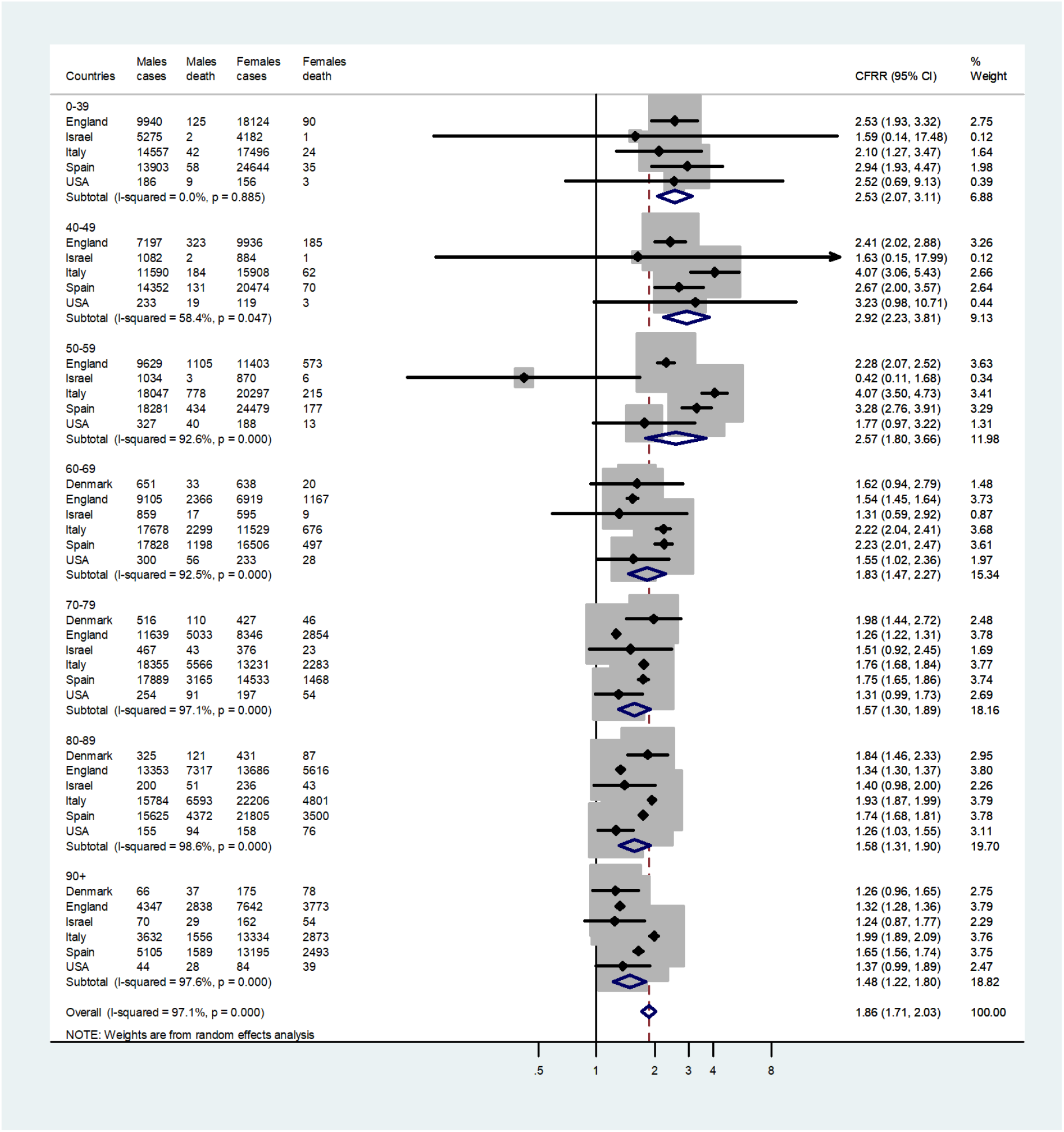
Forest plot of the ratio of the male to female CFRs in six countries for each age group.

### Sensitivity analysis

To evaluate the effect of individual countries and years on the pooled CFRs, we performed leave-one-out sensitivity analysis and recomputed the pooled CFRs. After omitting one country at a time, the pooled CFRs remained very similar. Similar results were obtained after omission of another age group at a time. Thus, no single country or age group substantially influenced the pooled CFRRs. This confirms that the results of this study are stable and robust.

### Meta-regression analysis

Meta-regression results are shown in Figure 2. They revealed that the age groups (p<0.0001) contributed to almost all the source of heterogeneity, with very little contributed by the differences between the countries (P=0.206).

**Figure 2.**
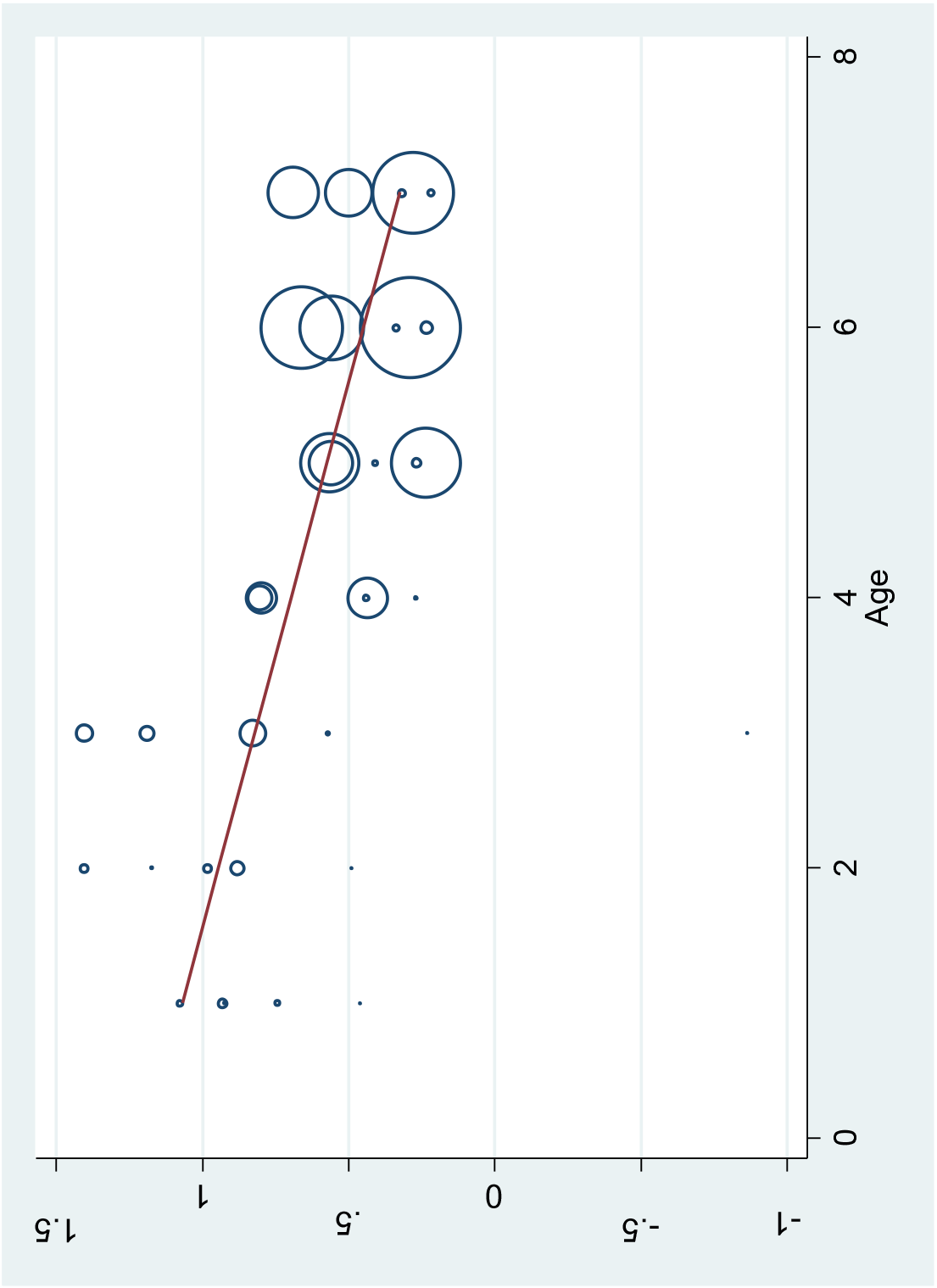
Meta regression analysis of the ratios of the male to female CFRs for each age group in six countries. Age groups: 1=0-39, 2=40-49, 3=50-59, 4=60-69, 5=70-79, 6=80-89, 7=90+

The age-related trend in the male to female CFRRs is shown in Figure 3. Generally, the CFRR increases slightly up to age 50, declines markedly until age 60, and then declines more gradually. Since the CFR is much lower under the age of 60, there are fewer deaths per cases and consequently the confidence intervals are wider.

**Figure 3:**
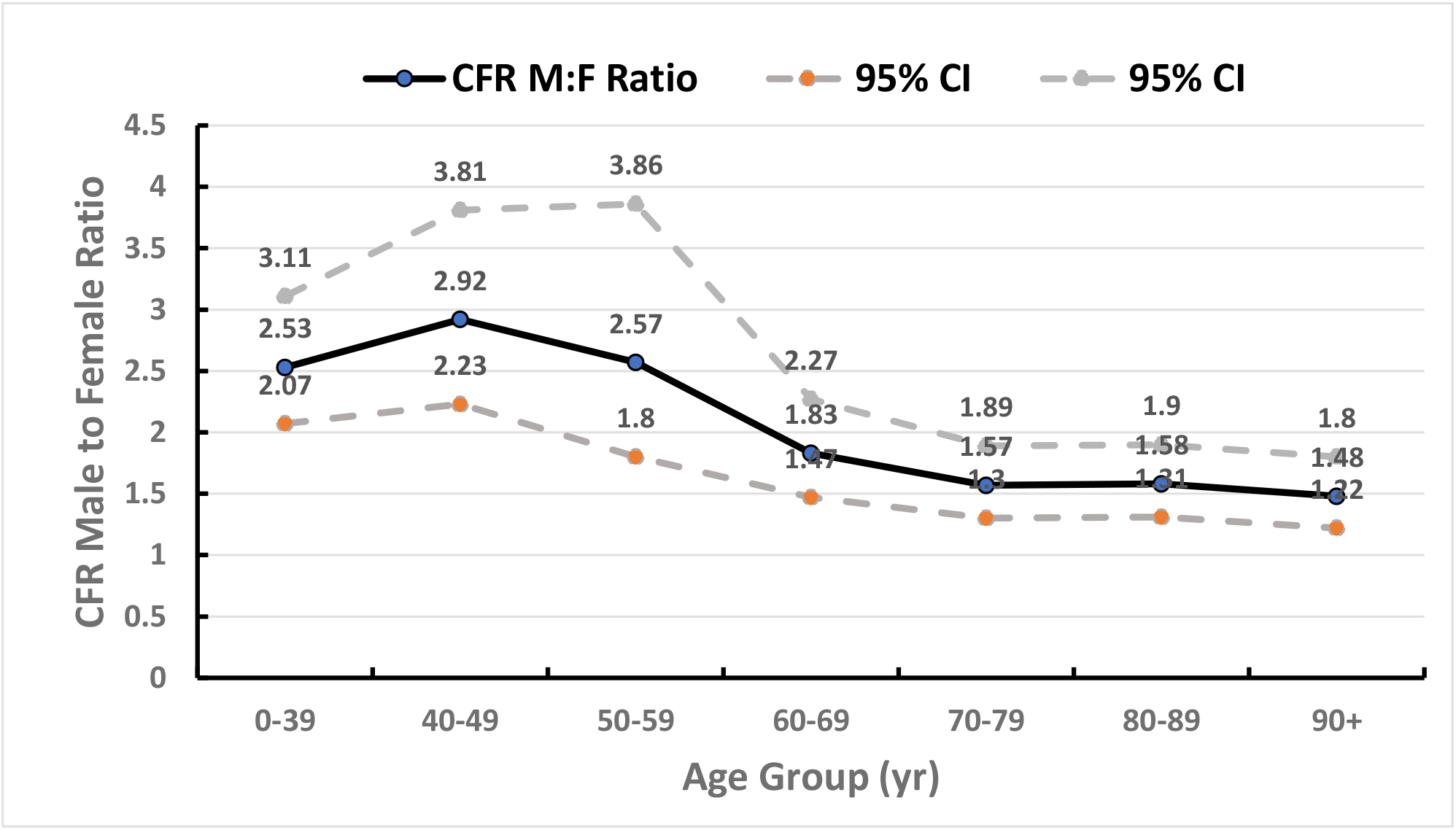
The pooled male to female CFRR by age group, pooled for six countries, together with the 95% CI lower and upper limits, derived from the meta-analysis.

### Discussion

Based on a meta-analytic study of national data from six countries, over a period of about two months, we found that the CFRs for COVID-19 were higher in males in all age groups and the sex ratio of the CFRs varied from 1.48 higher in males in the age over 90, to 2.92 in the age 40-49. The excess case-fatality in males in each age group was remarkably consistent over different countries. Our findings provide evidence of a large and consistent excess CFR in males for COVID-19, for all age groups, over six countries. This study supports and extends the findings in other studies (4, 5).

A major strength of the study is that it is based on national data with large populations and a large number of deaths and cases, based on reliable denominators. Selection bias has been minimized by using national data, which should be representative of each country. The inclusion of six countries, allowed us to evaluate the consistency of the findings over different populations. We do not believe that excluding countries that have poor diagnostic facilities or reporting has created an important source of selection bias which would affect the sex differences in CFRs. The calculation of the CFR can be affected by a number of potential biases. Selection bias is clearly present when calculating the denominator on the basis of reported cases. If only those with more severe symptoms are tested this will affect the denominator of the CFR and will depend on the testing strategy of each country. For example, if tests are restricted to the more severe cases, and if females tend to have a less severe disease this would selectively underestimate the number in the denominator for the female CFR and inflate the female CFR. Selection bias may also affect the numerator if only deaths occurring in hospital are reported but should not differ between the sexes.

An alternative measure of the severity of the disease is the infection fatality rate. The denominator of the infection fatality rate (IFR) includes both symptomatic and asymptomatic cases, detected either by active screening or incidentally during investigations of outbreaks. Since the IFR is rarely available for COVID-19, in this paper we examined the sex differences in the case-fatality rates (CFR), where the denominator is only reported diagnosed cases. Although the CFR is likely to be substantially higher than then the IFR, due to the smaller denominators, there is no reason to suspect that the sex ratio of the CFR will be substantially different from the sex ratio of the IFR.

Information bias can be present in both the numerator and denominator of the CFR. The definition of the cases may be biased due to the variability of the sensitivity and specificity of the diagnosis of COVID-19. Information bias in the numerator can occur when the cause of death is coded. This could be particularly problematic in elderly people with multiple co-morbidities. This could also differ between the sexes due to differences in comorbidities. Since the clinical manifestations of COVID-19 vary widely, there could be significant differences in case definitions and allocation of causes of death. However there is no reason to suspect that the reporting is related to the sex of the patient. Finally, differences that may exist in the laboratory methods used to confirm infection should also be unrelated to the sex of the patient. There is a lag time in the occurrence of the deaths relative to the number of cases reported. Thus the cases in the denominator are usually an overestimate of the true denominator which should be the number of cases reported sometime earlier (10, 11).

However, this should not be different between the sexes unless males progress to death more rapidly than females. In this case the CFR would be inflated in males compared with females. While this study cannot provide exact description of the mechanisms in the sex differences observed, it is possible to postulate some explanations. Regarding possible cultural factors, in the countries in this study, there is no evidence that the sex of the patient influenced the medical care given for COVID-19. Similarly, there is no evidence to suggest that in these countries, adult men are more likely than women to delay medical care for acute conditions of comparable severity, although in general there is evidence that women are more likely to seek medical care (17). Sex differences in exposure due to behavioural factors could play a part in the incidence of the cases. However, since this study focuses on CFRs that should not influence the result. The severity of COVID-19 has been shown to be strongly associated with underlying conditions in the patients (18). These include hypertension, diabetes, and obesity. However, in a large study in the United Kingdom (19) males were still at higher risk of death after controlling for co-morbidities.

The genetic and hormonal differences between males and females have been suggested as possible explanations for the higher case-fatality in males (4). The SARS-CoV-2 virus uses the ACE2 receptor to enter the cells. It has been reported that circulating ACE2 levels are increased in male compared with female subjects, in patients with diabetes or cardiovascular diseases, and in prostate epithelial cells (20, 21). The different hormonal environment could have extended pathophysiological role in SARS-CoV-2 occurrence, with testosterone, causing men to develop more serious complications related to the SARS-CoV-2 infection (22). In general, for SARS-CoV, it has been reported, that estrogen signalling in females may directly suppress SARS-CoV replication via effects on cellular metabolism (23).

Estradiol promotes innate immune signalling pathways, including macrophages, dendritic cells (DCs), granulocytes, and lymphocytes cell development. The hormone also enhances production of pro-inflammatory cytokines and chemokines in response to TLR ligand stimulation of dendritic cells and macrophages, a phenomenon that may explain the superior immune response to infection in pre-menopausal females (24-26). In COVID-19 infected women the production of inflammatory IL-6 (one of the main components of cytokine storm) is lower than in males and is often correlated with a better longevity (27). Testosterone has the effect of depressing the innate and adaptive immune response (6, 28). Thus, it is conceivable that sex hormones are implicated in the mechanism of infection by COVID-19.

However, sex hormone involvement in ACE2 regulation is likely to be important under the age of 50, when differences in hormone levels between men and women are significant (29-31). At older ages, after the age of 50-60, hormone differences probably have less significance because of profound changes in the hormonal milieu in both women and men. Although estrogen concentrations in men are about 200 times lower than that of testosterone, over the age of 50, they are higher than the concentrations in postmenopausal women (32). In this study, the male excess in CFR’s was evident at all ages, but somewhat lower with increasing age. This could be due to a reduced effect of the differences in sex hormones.

The severity of SARS-CoV-2 appears to depend on the interaction between the virus and the individual’s immune system differentially by sex and age (33). Genetic differences between males and females could be relevant at all ages. Females generally exhibit elevated humoral and cell-mediated immune responses compared to males (6). X chromosomes contain genes associated with the immune system and the presence of two X chromosomes plays a major role in enhancing both innate and adaptive immune responses. Genetic factors could play a part through an interaction with sex hormones (34). The ACE2 and Ang-II receptor type 2 gene are both located on the X-chromosome and this may impact male susceptibility to COVID-19. X-chromosome genes could encourage mosaic advantage in females and sexual dimorphism that might mitigate viral infection and inflammation due to cytokine storms (35). At older ages, genetic factors could be more dominant. With ageing, sex chromosomes undergo changes that influence their possible contribution to risk for diseases (36).

In conclusion, the remarkably consistent excess COVID-19 CFRs in males in all countries and in all age groups, suggests that sex-specific factors influence the severity of COVID-19. The substantially higher CFR in the males in the younger age groups suggest that a hormonal factor may be operating, whereas at older ages, genetic differences are likely to dominate. These findings should stimulate research on sex as a biological variable in the pathogenesis of COVID-19.

## Data Availability

All the data are freely available to the public

